# Accuracy and efficiency of using artificial intelligence for data extraction in systematic reviews. A noninferiority study within reviews

**DOI:** 10.64898/2026.02.25.26347053

**Authors:** Daniel C.W. Lee, Kate M O’Brien, Justin Presseau, Serene Yoong, Christophe Lecathelinais, Luke Wolfenden, James Thomas, Anneliese Arno, Brian Hutton, Rebecca K Hodder

## Abstract

**Background:** Systematic reviews are important for informing public health policies and program selection; however, they are time- and resource-intensive. Artificial intelligence (AI) offers a solution to reduce these labour-intensive requirements for various aspects of systematic review production, including data extraction. To date, there is limited robust evidence evaluating the accuracy and efficiency of AI for data extraction. This study within a review (SWAR) aimed to determine whether human data extraction assisted by an AI research assistant (Elicit^®^) is noninferior to human-only data extraction in terms of accuracy (i.e. agreement) and time-to-completion. Secondary aims included comparing error types and costs.

**Methods:** A two-arm noninferiority SWAR was conducted to compare AI-assisted and human-only data extraction from 50 RCTs chronic disease interventions. Participants were randomised to extract all data required for conducting a review, using either the AI-assisted or human-only method. Accuracy was assessed using a three-point rubric by an independent assessor blinded to group allocation, based on agreement between extracted data and the assessor. Accuracy scores were standardized to a 0–100 scale. Analysis included overall and subgroup accuracy (data group and data type) using paired t-tests. Time-to-completion was self-reported by data extractors. Type of errors were coded by type and severity, and costs were calculated for data extraction, preparation of files, training and the Elicit^®^ Pro subscription.

**Results:** There was no difference in overall accuracy between the AI-assisted and human-only arms (mean difference (MD) 0.57 (on a 0-100 scale), 95% confidence interval (CI) -1.29, 2.43). Subgroup analysis by data group found AI-assisted to be more accurate than human-only data extraction for data variables describing ‘intervention and control group’ (MD 4.75, 95% CI 2.13, 7.38), but otherwise no subgroup differences were observed. AI-assisted data extraction was significantly faster (MD 24.82 mins, 95% CI 18.80, 30.84). The AI-assisted arm made similar error types (missed or omitted data: AI-assisted 3.6%, human-only 3.4%) and severity (minor errors: AI-assisted 6.7%, human-only 6.5%) and cost $181.98 less than the human-only data extraction across the 50 studies.

**Conclusion:** AI-assisted data extraction using Elicit^®^ showed noninferior accuracy, faster completion times, similar error types and severity, and lower costs compared to human-only extraction. These efficiency gains, without loss in accuracy suggest AI-assisted data extraction can replace one human-only data extractor in future systematic reviews of RCTs. Future research should explore different models of AI data extraction such as two AI-assisted extractors or AI-only extractor with human-only extractor, and comparison of AI-assisted to AI-only.

## Background

Systematic reviews are critical for informing public health policies ^(1, 2)^, but data extraction remains one of the most labour-intensive and error-prone step, with error rates up to 50% ^(3, 4)^. Best practice recommends that two authors independently extract data, followed by a consensus process to resolve any discrepancies, or by involving a third authors when consensus cannot be reached ^(5)^. Tools like Excel ^(6)^, Word ^(7)^ and Covidence ^(8)^ are commonly used to improve efficiency, yet data extraction can average 107 minutes per study per authors ^(9, 10)^, increasing to 172 minutes with consensus ^(10)^, and potentially longer for complex reviews or inexperienced authors ^(3)^. Accuracy in data extraction is particularly important as errors can seriously affect the validity of the review findings as authors rely on extracted data for analyses (e.g. meta-analysis), drawing conclusions and making recommendations ^(11)^. Given the need for timely and well conducted systematic review evidence to inform decision making, it is important to investigate innovative methods that can improve both accuracy and efficiency of data extraction for systematic reviews.

Artificial intelligence (AI) automation, particularly generative AI (GenAI) ^(12)^, offers a promising solution to address the growing challenges of maintaining up-to-date, high-quality systematic reviews without requiring programming expertise ^(13-15)^. To date, AI automation tools have been developed and tested across various systematic review stages, including literature searches, screening abstracts and full-text articles, extracting data, and risk of bias assessment ^(16-21)^. Such tools can fully automate (no human interaction) or partly automate (includes human interaction) the systematic review workflow reducing time and burden associated with these processes ^(22)^. As the use of GenAI automation tools presents new opportunities, various global evidence synthesis organisations have developed guidelines for its responsible use ^(23-26)^. For example, the ‘Responsible AI in Evidence SynthEsis’ (RAISE) guidance document encourages evaluation and validation studies to inform best practice guidance and foster collaboration with tool developers, to ensure tools are fit-for-purpose ^(25)^.

Given the rapid evolution of AI, rigorous evaluation of new GenAI automation tools is essential. Emerging evidence suggests that GenAI is capable of producing accurate data extraction. For example, a non-randomised study within a review (SWAR) evaluated semi-automated data extraction of six ongoing reviews with both randomised controlled trials (RCTs) and non-RCTs, and found that the use of AI-assisted data extraction using Claude 2 achieved an overall accuracy of 91.0%, where accuracy was defined as agreement with two blinded human assessors, and had fewer incorrect extractions (9.0% vs. 11.0%) when compared with human-only extractors ^(27)^. Similar levels of accuracy or acceptable quality have been reported by other studies, though these often employed less robust designs such as proof-of-concept, lack of blinding and limited training of outcome assessors ^(28-30)^. GenAI also shows potential for improving efficiency. Several studies comparing GenAI with human-only data extraction reporting time savings. For instance, the non-randomised SWAR highlighted above found AI-assisted extraction saved a median of 41 minutes per study when compared to human-only methods (84 vs. 125 minutes) ^(27)^. Despite the promise of GenAI, examination of the benefits of automated and/or semi-automated data extraction requires further investigation ^(31)^, as evident from a 2025 living systematic review that found few accessible tools ^(14)^.

One widely available GenAI tool developed for data extraction is Elicit^®^. Elicit^®^ uses LLMs to extract structured information from PDFs or online studies and presents them with quotes from the source enabling user to verify and amend the output ^(32)^. Although evaluations of Elicit^®^ remains limited, four studies have assessed its data extraction capabilities, reporting high accuracy, and comparable or superior comprehensiveness relative to human extractors ^(29, 33-35)^. However, these studies focused on fully automated extraction and assessed only a small number of variables. Cost is another important consideration, particularly given the limited funding available to many researchers ^(36)^. Elicit^®^ offers both free and premium options ^(37)^. Of the four studies, only one considered cost, without evaluating whether time savings translated into overall cost savings ^(35)^. Therefore, a comprehensive evaluation is needed to assess Elicit^®^’s performance as an AI-assisted extractor across a broader range of data variables, and to account for both direct costs and the potential cost implications from improved efficiency.

Given the central role of systematic reviews in informing policy and practice, it is essential to establish whether GenAI-assisted extraction achieves accuracy comparable to human-only extraction while offering potential efficiency gains. A noninferiority trial is an appropriate (though rarely used) study design for this purpose as it is designed to test whether a new method (i.e. GenAI-assisted extraction) is not meaningfully worse than the reference standard (i.e. human-only extraction) within a predetermined acceptable margin ^(38)^. Therefore, on the basis of the evidence gap, we conducted a noninferiority SWAR to evaluate the accuracy and time efficiency of using Elicit^®^ for GenAI-assisted data extraction compared with human-only extraction. For brevity, we refer to GenAI-assisted data extraction as AI-assisted data extraction from this point forward.

### Aims of the trial

This noninferiority SWAR aimed to determine whether human data extraction assisted by an AI research assistant (Elicit^®^) is noninferior to human-only data extraction in terms of accuracy and time-to-completion. Secondary aims included comparing error types and costs.

## Methods

This trial adheres to the CONSORT guidelines for reporting noninferiority and equivalence randomised trials^(38)^. The protocol was prospectively developed and retrospectively registered, available from osf.io/uykg8. Approval to conduct this trial to the methods described below was obtained from the University of Newcastle’s (UON) Human Research Ethics Committee, H-2024-0320.

### Trial design

A two arm, parallel-group, noninferiority SWAR ^(39)^ was conducted to test AI-assisted and human-only methods for extracting data from RCTs. The RCTs were sourced from a 2022 update of a Cochrane systematic review that examined the effectiveness of obesity prevention interventions on the weight of children aged 6-18 years ^(40)^.

### Participants and studies

#### Eligibility

Graduate students or post-doctoral researchers with basic knowledge in the research area (i.e. chronic disease prevention in children and adolescents), sufficient English literacy, and willingness to complete all assigned extraction were eligible to participate. Prior experience in systematic review data extraction was preferred but not required. Participants were not required to have any prior experience with data extraction using GenAI or Elicit^®^. Participants were ineligible if they previously worked on the original systematic review ^(40)^. Studies were eligible if they were school-based RCTs, written in English, with a readable portable document format (PDF) processable by Elicit^®^.

#### Recruitment

Invitations to participate in the trial were sent via existing email distribution lists of the research team. The two most experienced participants were recruited and randomly allocated across the two trial arms. Both participants were reimbursed as research assistants for their participation.

### Data extraction

Fifty-one of the 58 variables for data extraction were drawn from Cochrane’s data extraction form template for RCTs ^(41)^. Seven were excluded from the trial for various reasons (e.g. only used for record keeping (e.g. Study ID)). Data variables were categorised into six data groups (general information, methods, participants, intervention and control groups, data and analysis and other information) and data type (numeric only, alphabetic only or both) (Appendix A).

Participants were provided with training on the expectations of data extraction and piloted the data extraction form with five unrelated studies (non-school-based) to ensure consistency and competency prior to completing data extraction. No training was provided on using Elicit^®^.

### Trial arms

#### AI-assisted arm

At the time of this trial, Elicit^®^ offered three tiers: Basic (free), Plus ($183.60 /year), and Pro ($763.70/year). The Pro subscription (December 2024 version) was used in this trial, which enabled ‘high-accuracy mode’ for all 50 included studies. This mode extracted data from tables and provided supporting quotes and reasonings to enhance accuracy ^(42)^. In the current version, high-accuracy mode is standard across all tiers^(37)^.

Elicit^®^ includes pre-existing prompts for various data variables from the Cochrane data extraction form (e.g. study design, region), which was used in this trial. For data variables without pre-existing prompts or lacked the required specificity, new custom prompts were developed by two authors (DL, RH). New prompts were drafted and assessed against non-school-based studies, previously extracted data and verified against the source PDFs. Prompts with poor accuracy were refined and retested until both authors (DL, RH) agreed with the output. Of the 51 variables, 29 prompts were generic to any review and 22 were review specific.

Appendix B presents the final prompts used.

Both participants received a link to Elicit^®^’s notebook (Figure 1) which contained extracted data, a secure OneDrive folder containing blank data extraction forms (Appendix C) and PDFs of all relevant records (e.g. primary outcome paper, associated protocols, trial registrations). Both participants were advised they were free to retain all, some or none of Elicit^®^’s output when completing data extraction. To reflect real-world use, participants accessed Elicit^®^ directly via a link, rather than pre-filled forms (Appendix D). Both participants were unable to modify prompts but could interact with Elicit^®^ to locate the origin of source quotes.

**Figure 1.**
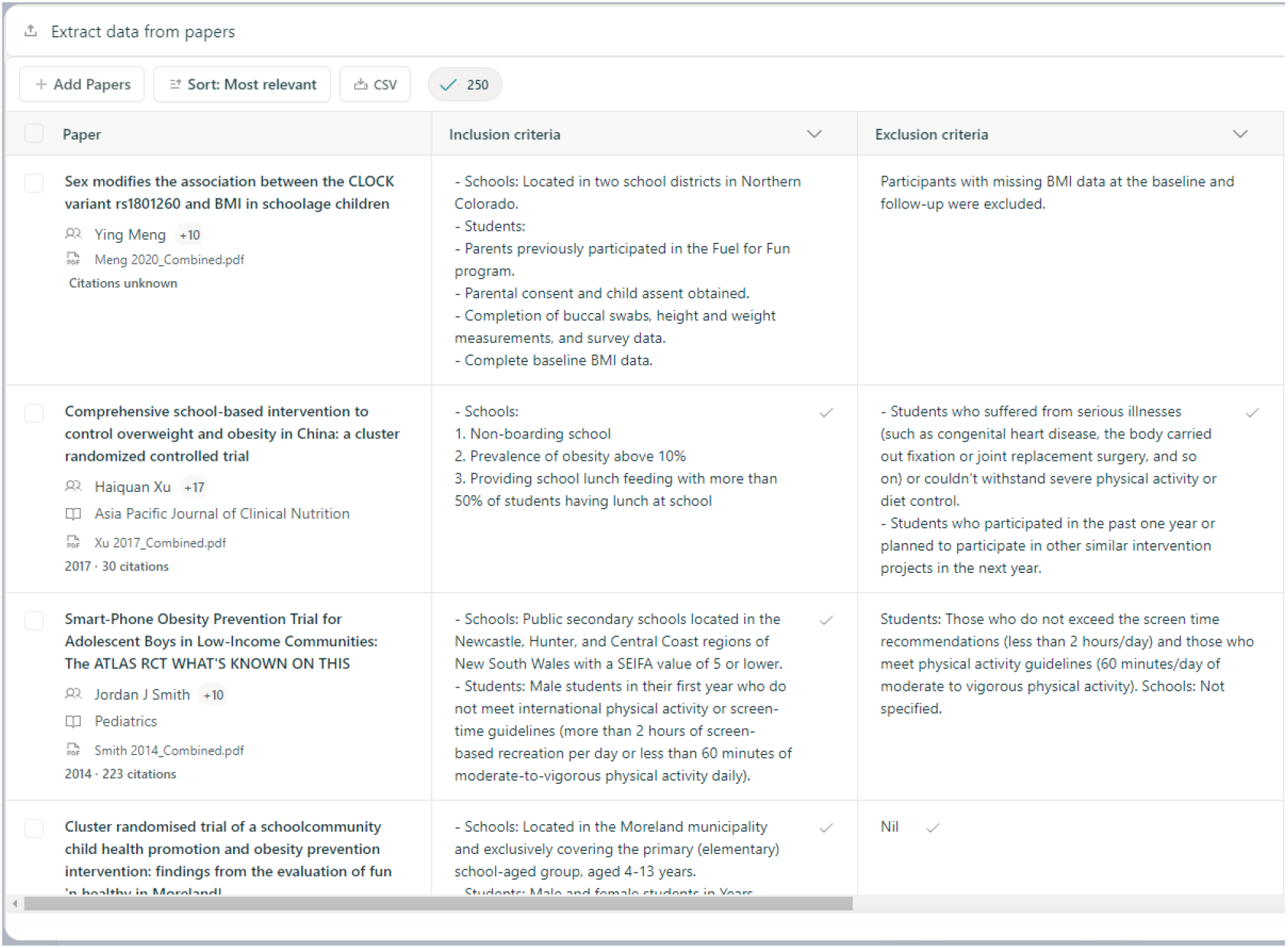
Example of Elicit^®^’s notebook

#### Human-only arm

Both participants were provided with a similar OneDrive folder as the AI-assisted arm excluding the link to Elicit^®^’s notebook and were advised to complete data extraction using only the PDFs available to them.

### Outcomes

#### Primary outcomes

##### Accuracy

To assess the accuracy of the data extracted for each item, a single blinded, post-doctoral researcher (KO), experienced in Cochrane data extraction, an author of the source review, and not involved in data extraction or prompt engineering, independently assessed the accuracy of each data variable across all studies using the source PDFs, a marking rubric and Cochrane extraction forms from the published review ^(40)^. The marking rubric was adapted from published resources ^(30, 43, 44)^ and piloted by the research team (Appendix D). The marking rubric outlined detailed accuracy criteria for each item scored on a three-point scale: 0 for incorrect, 1 for partially correct and 2 for correct. See Table 1 for more information. Of the 51 data variables, 41 were scored either 0, 1 or 2 and 10 were scored either 0 or 2 (due to their simplicity (e.g. study design, unit of allocation, trial registry number), with a maximum possible score of 102. The final set of correct answers (i.e. gold-standard) are provided in Appendix G.

**Table 1:** Example of marking rubrics.

| Data points | 0 (Incorrect) | 1 (Partially correct) | 2 (Correct) |
| --- | --- | --- | --- |
| **Trial registration number** (if available) | Missing, fabricated, or completely incorrect information about the trial registration or protocol. | Not applicable. | Accurately extracts trial registration numbers and protocol citation details, matching the source perfectly. |
| **Setting** (including location (city, state, country)) | Missing, vague, or incorrect description of the setting (e.g., does not mention location or context). If the study reports vaguely, do not mark as incorrect. | Partially correct; includes the location (e.g., city) but lacks full social context or omits specific details.<br>If any part of the location are missed.<br>E.g. Nunoa, Santiago, Chile but only reports Santiago, Chile | Fully describes the setting, including detailed information about location (e.g., city, state, country) and social context (e.g., rural/urban, community type). Mark correct if consistent with what is reported in the study. |

##### Time-to-complete

To measure efficiency, time-to-complete was measured as the total time spent on all tasks necessary for preparation and data extraction. Time taken for preparation were inclusive of template editing, file organisation, piloting, prompt engineering, uploading PDFs, interacting with Elicit^®^. To record the time taken to complete data extraction, participants were instructed to self-record the start time and end time of data extraction for all study, including the any breaks taken in between, in a monitoring form in Excel ^(6)^.

#### Secondary outcomes

##### Types and severity of error

The lead author (DL) coded the type (missed or omitted data, misallocated data, incomplete data, false data, incorrect calculations/inferences, or other) and severity of each error (major error, minor error, or inconsequential error) for scores of 0 or 1 (Table 2) using categories adapted from previous publications ^(27,30)^.

**Table 2:** Types of errors and severity of error for data extraction.

| Type of error | Definitions |
| --- | --- |
| Missed or omitted data | Data that were available in the reference standard (or source PDF) but were either missed or omitted. |
| Misallocated data | Data that were available in the reference standard (or source PDF) but were allocated to the wrong data extraction item or taken from the wrong source PDF |
| Incomplete data | Relevant and correct data provided but missing key details |
| False data (“Hallucination”) | Fabricated data that seem to be generated by the AI or data extractor |
| Incorrect calculations/inferences | Mathematically incorrect calculations of data elements based on information provided in the source study report, including rounding errors. Includes wrongly drawn answers such as CRCT but reporting individuals as unit of allocation |
| Other | Optional “other” field if none of the above apply, for example typos or misunderstandings of instructions |
| **Severity of error** |  |
| Major error | This error significantly compromises the accuracy of the data, and, if uncorrected, could lead to erroneous conclusions; for example, grossly incorrect calculations or misallocated data. |
| Minor error | This error is less severe than a major error but still impacts the quality of the existing data; for example, small calculation errors or rounding errors that do not critically affect the data's overall utility. |
| Inconsequential error | This difference most likely would not impact the interpretation of the data, for example differences in the level of detail of inclusion criteria |

Adapted from Gartlehner 2024 and 2025 <sup>(27, 30)</sup>.

**Table 3:** Mean time-to-complete (minutes) by participants.

|  | AI-assisted arm (mins) | Human-only arm (mins) | Total mean (mins) |
| --- | --- | --- | --- |
| Participant A | 29.7 | 73.6 | 51.7 |
| Participant B | 18.0 | 53.9 | 35.9 |

##### Costs

Cost for each intervention arm included research assistant labour remuneration ^(45)^ (including data extraction, time spent in file preparation, setting up the AI-assisted arm, training) and annual Elicit^®^ Pro subscription fee ($763.70 per year) ^(37)^. The cost of Elicit^®^ Pro subscription was adjusted to reflect the actual days of use from the commencement of prompt engineering to the completion of data extraction. All costs are reported in Australian Dollars (AUD).

### Noninferiority margin and sample size

The noninferiority margin was set at 10% based on a prior study that evaluated the noninferiority of a risk of bias AI tool ^(46)^, the only noninferiority trial on GenAI automation tool identified by the authors. Notably, this margin aligns with the stricter thresholds commonly used in noninferiority assessments of antibiotics ^(47-49)^. This meant that for accuracy, the AI-assisted arm would be considered noninferior to the human-only arm if on average, less than 10% of items per study were different (n=51), which is equivalent to a difference in score of 5.1 to 10.2 or a score of 5 to 10 when standardised to a 0-100 scale. For time-to-complete, the AI-assisted arm would be considered noninferior to the human-only arm if on average, less than 10% of time-to-complete was different to the human-only arm.

Sample calculation was completed using PS ^(50)^. Sample size was calculated based on the power function of paired t-test. For the calculations, we used 50 pairs of studies, assumed a type I error rate of 2.5% (a convention for noninferiority studies) ^(50)^, 90% power and unknown SD on the difference which resulted in a detectable difference of 0.51 z-score units.

### Trial procedures

#### Sequence generation

Of the 195 RCTs included in the source review ^(40)^, 74 were ineligible (71 not school-based, three not written in English). All studies were had readable PDFs. Of the remaining 121 eligible RCTs, 50 were randomly selected and used in each trial arm. Within each arm, participants were randomly allocated to extract data using either the AI-assisted or human-only method for each of the 50 studies (Figure 2) based on a list of randomised numbers. The list was generated by an independent statistician (CL), using a random number function in Microsoft Excel ^(6)^ with participants allocated in a 1:1 ratio to each study arm.

**Figure 2.**
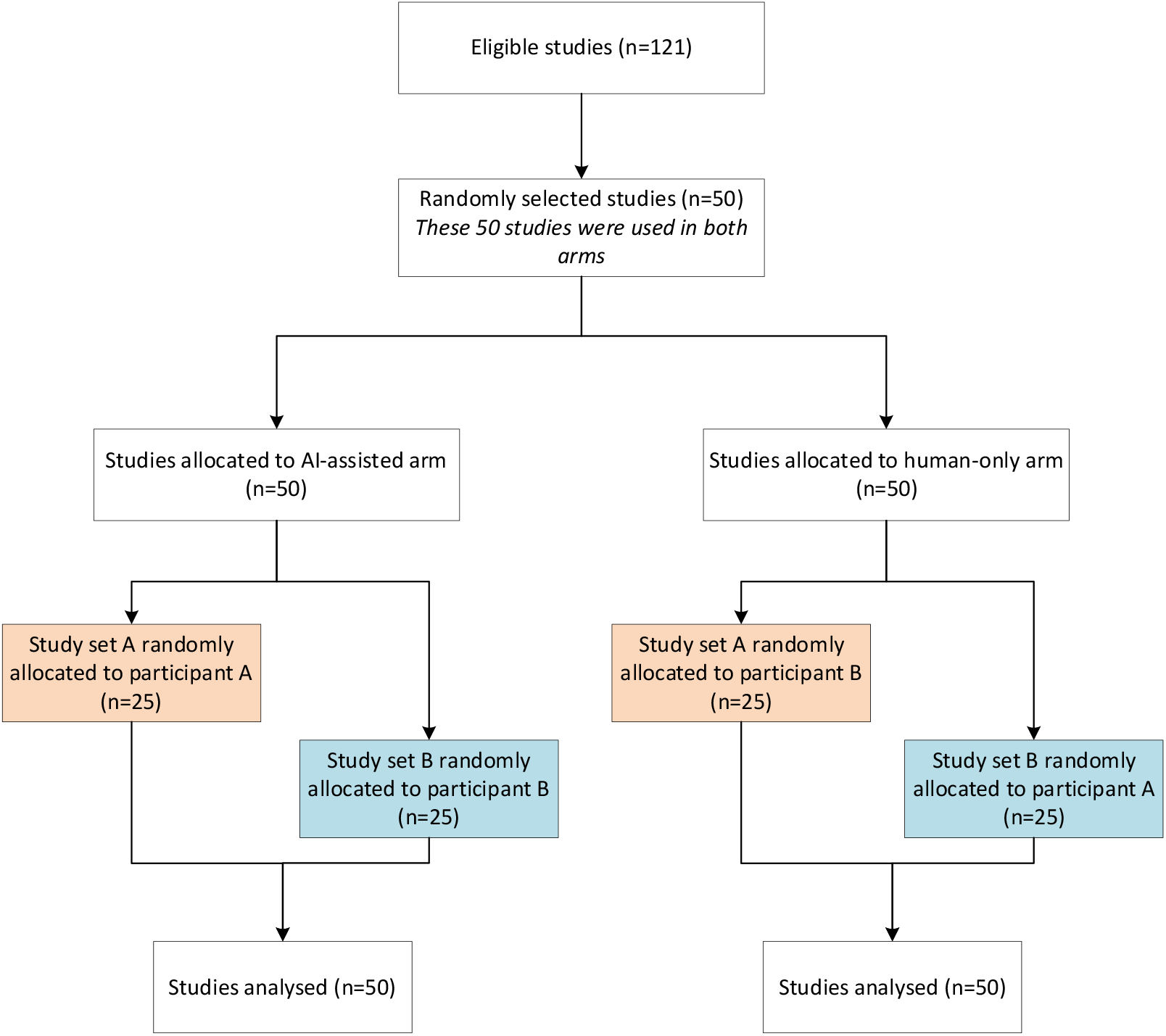
Study flow diagram

Participants were listed alphabetically by surname and assigned the label “Participant 1” or “Participant 2” accordingly. Studies were then randomly allocated to the AI-assisted arm and assigned to one of the two participants. The other participant, who did not receive the AI-assisted version of a given study, was assigned the same study in the human-only arm.

#### Allocation concealment

The lead author was not blinded to the allocation sequence.

#### Blinding

Only the participant and the lead author (DL) had access to the OneDrive folder to maintain blinding between participants. Participants knew their group allocation, as full blinding was not possible.

Upon completion of data extraction in each arm, the DL de-identified all forms by assigning unique trial numbers and standardising formatting (e.g. font type and sizes). The accuracy assessor (KO) was blinded to study arms and independently assessed the accuracy of each data variable. All analyses were conducted by RH, who remained blinded to group assignments.

### Statistical analysis

Participant characteristics and costs were reported using descriptive statistics. The unit of analysis was the individual study. Accuracy analysis was conducted in SAS, version 9.4 ^(51)^ and time-to-complete analysis was conducted in Microsoft Excel, Version 2504 ^(52)^. Normality of the data was assessed through visual inspection of histograms.

#### Accuracy

Accuracy scores were reported using descriptive statistics and standardised to a 0-100 scale. The scores of each data variable within each study were summed to provide a total accuracy score per study. A difference score was calculated for each study by subtracting the AI-assisted arm by the human-only arm, resulting in a set of 50 paired difference scores. A paired t-test was then applied to these 50 difference scores. Noninferiority was assessed by comparing if the differences between both arms was within 10% and within the upper and lower 95% confidence intervals (CI) of the mean difference, with a significance level of α=0.025.

Accuracy was examined within subgroups categorised by data group (i.e. methods, participants, intervention and control group, data and analysis and other information) and data type (i.e. alpha, numerical, both) using a t-test where more than one variable was available.

#### Time-to-complete

To assess the impact of the AI-assisted approach compared to the human-only approach, a difference score was calculated for each study by subtracting the total time for the AI-assisted arm from the total time for the human-only arm. A paired t-test of difference scores was conducted to examine time differences.

Noninferiority of the AI-assisted compared to the human-only approach was assessed by comparing if the differences between both arms was within 10% and within the upper and lower bound of the 95% confidence interval (CI) of the mean difference, using a two-tailed significance level of α=0.025.

A sensitivity analysis was also conducted to compare the time-to-complete between both arms exclusive of the additional of preparation time for both arms. Time taken to prepare both arms was calculated by averaging the time spent on each task (e.g. editing template, file organisation and piloting) for each study. The AI-assisted arm included additional tasks specific to the arm, specifically, prompt engineering, uploading PDFs, interacting with Elicit^®^, and running the data extraction.

#### Types and severity of error

Descriptive statistics were used to describe the types and severity of errors.

#### Costs

Descriptive statistics were used to describe the breakdown of costs by trial arms.

## Results

Visual inspection of histograms suggested no evidence of non-normality. Two participants undertook data extraction between December 2024 and January 2025. Both participants held master’s degrees, had prior experience as research assistants, and familiar with data extraction. One had led a systematic review. Neither had prior experience using Elicit^®^. The mean time-to-complete by participants had a difference of 15.8 minutes (Table 5).

### Accuracy

The overall accuracy mean difference between arms was 0.57 (95% CI -1.29, 2.43). Both the mean difference and the noninferiority bound of the 95% CI were within the predefined inferiority threshold of less than 10 points difference (Figure 3). The mean overall accuracy score for the AI-assisted and human-only data extraction arms were 85.8 (range 74 - 99) and 85.3 (range 72 – 94) respectively. Mean accuracy score by data variable can be found in Appendix E.

**Figure 3.**
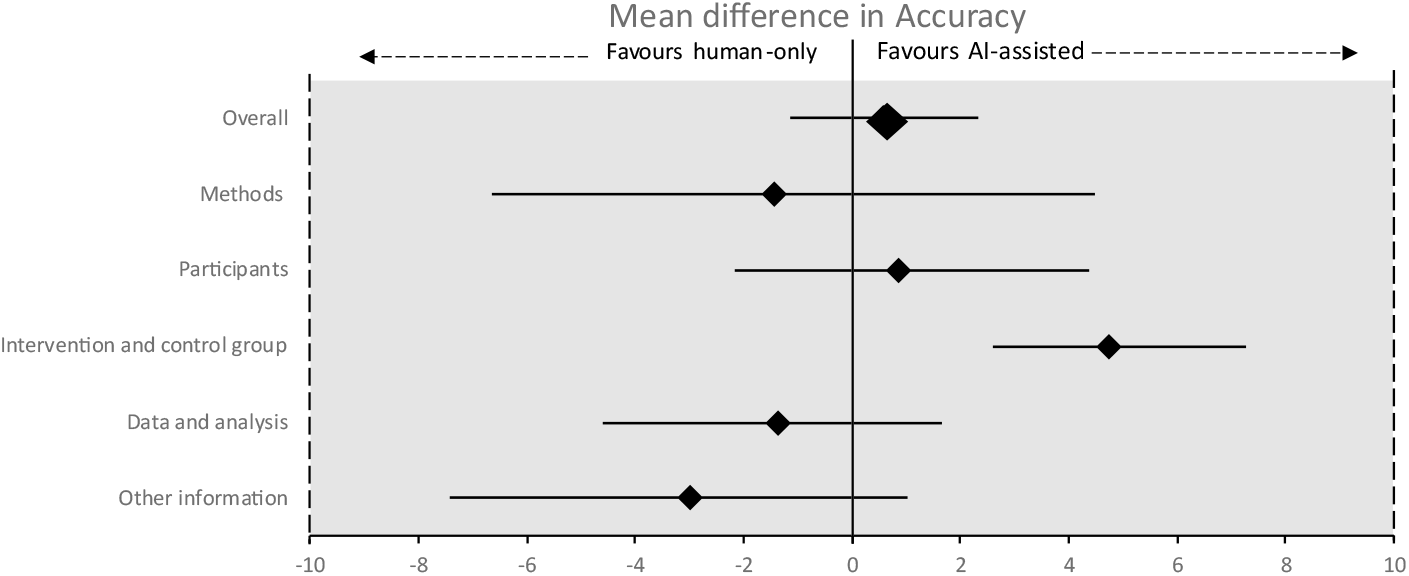
Overall and data group level accuracy results

Subgroup analysis by data group found no differences between all data groups except for *intervention and control group* (mean difference (MD) 4.75, 95% CI 2.13, 7.38; Table 4). Subgroup analysis by data type (i.e., alpha, numeric, both) found no differences between all data type ranged (Table 4)

**Table 4:**
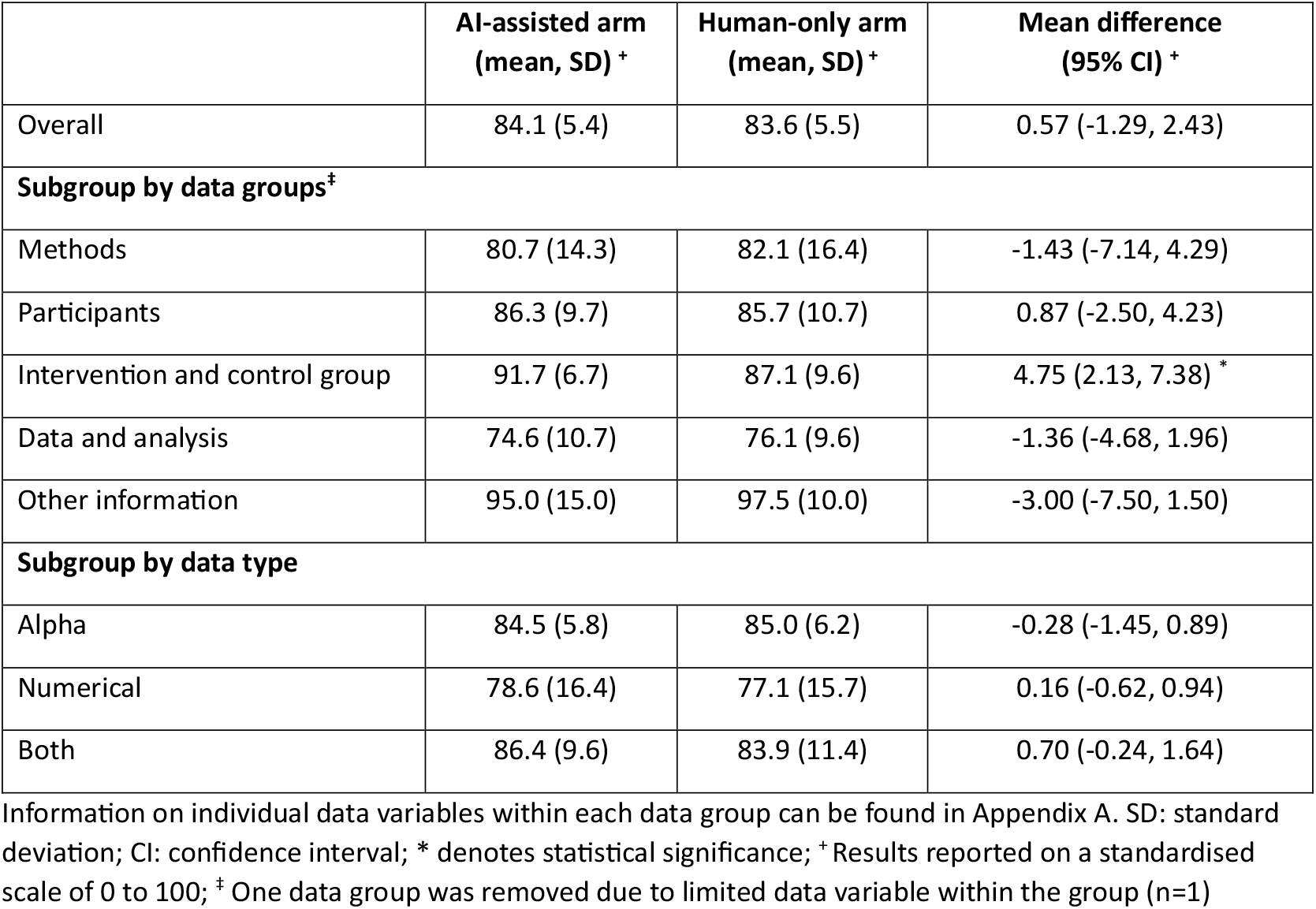
Accuracy of AI-assisted and human-only data extraction arms (n=50)

|  | AI-assisted arm<br>(mean, SD) <sup>+</sup> | Human-only arm<br>(mean, SD) <sup>+</sup> | Mean difference<br>(95% CI) <sup>+</sup> |
| --- | --- | --- | --- |
| Overall | 84.1 (5.4) | 83.6 (5.5) | 0.57 (-1.29, 2.43) |
| **Subgroup by data groups<sup>‡</sup>** |  |  |  |
| Methods | 80.7 (14.3) | 82.1 (16.4) | -1.43 (-7.14, 4.29) |
| Participants | 86.3 (9.7) | 85.7 (10.7) | 0.87 (-2.50, 4.23) |
| Intervention and control group | 91.7 (6.7) | 87.1 (9.6) | 4.75 (2.13, 7.38) * |
| Data and analysis | 74.6 (10.7) | 76.1 (9.6) | -1.36 (-4.68, 1.96) |
| Other information | 95.0 (15.0) | 97.5 (10.0) | -3.00 (-7.50, 1.50) |
| **Subgroup by data type** |  |  |  |
| Alpha | 84.5 (5.8) | 85.0 (6.2) | -0.28 (-1.45, 0.89) |
| Numerical | 78.6 (16.4) | 77.1 (15.7) | 0.16 (-0.62, 0.94) |
| Both | 86.4 (9.6) | 83.9 (11.4) | 0.70 (-0.24, 1.64) |

Information on individual data variables within each data group can be found in Appendix A. SD: standard deviation; CI: confidence interval; \* denotes statistical significance; <sup>+</sup> Results reported on a standardised scale of 0 to 100; <sup>‡</sup> One data group was removed due to limited data variable within the group (n=1)

**Table 5:** Time-to-complete of AI-assisted and human-only data extraction arms.

|  | AI-assisted arm<br>(mean, SD) | Human-only arm<br>(mean, SD) | Mean difference<br>(95% CI) |
| --- | --- | --- | --- |
| Time-to-complete, with prep (mins) <sup>+</sup> | 48.5 (8.9) | 73.3 (16.7) | 24.8 (18.8 - 30.8) * |
| Time-to-complete, without prep (mins) | 23.8 (8.9) | 63.8 (16.7) | 39.9 (33.9 - 45.9) * |

SD: standard deviation; CI: confidence interval; \* denotes statistically significant; <sup>+</sup> Preparation tasks included template editing, file organisation and piloting. The AI-assisted arm also involved prompt engineering, uploading PDFs, interacting with Elicit®, and running data extraction.

Diamonds represent mean difference, and whiskers represent 95% confidence interval. AI: artificial intelligence. Overall: 0.57, 95% CI -1.29, 2.43; Methods: -1.43, 95% CI -7.14, 4.29; Participants: 0.87, 95% CI -2.50, 4.23;Intervention and control group: 4.75, 95% CI 2.13, 7.38; Data and analysis: -1.36, 95% CI -4.68, 1.96; Other information: -3.00, 95% CI -7.50, 1.50.

### Time-to-complete

The mean time-to-complete difference between arms was 24.8 minutes (95% CI 18.8, 30.8; Table 4). Both the mean difference and the noninferiority bound of the 95% CI were outside the noninferiority threshold of less than 10% difference (equivalent to <7.3 minutes), in the direction favouring the AI-assisted arm. The mean time-to-complete data extraction of the AI-assisted arm was also significantly faster on average than the human-only arm.

### Types and severity of error

Among the 5100 extracted data variables, 1007 errors were identified in both arms (AI-assisted 9.7%; human-only 10.1%). Of the 1007 errors, the most common types of errors for both arms were *missed or omitted data* (AI-assisted 18.5%; human-only 17.4%) and *incomplete data* (AI-assisted 17.4%; human-only 17.3%). False data was rare in both arms (AI-assisted 1.0%; human-only 1.0%). Overall, the most common severity of error for both arms were *minor error* (AI-assisted 34.1%; human-only 33.1%) (Table 6).

**Table 6:** Types and frequency of errors and severity.

|  | Number and percentage of errors of all data variables (n=1007) |  |  |  |  |  |  |  |
| --- | --- | --- | --- | --- | --- | --- | --- | --- |
|  | AI-assisted arm |  |  |  | Human-only arm |  |  |  |
| Severity of error<br>Error type | Major | Minor | Inconsequential | Total | Major | Minor | Inconsequential | Total |
| Missed or omitted data | 10<br>(1%) | 167<br>(16.6%) | 9<br>(0.9%) | 186<br>(18.5%) | 23<br>(2.3%) | 129<br>(12.8%) | 23<br>(2.3%) | 175<br>(17.4%) |
| Misallocated data | 2<br>(0.2%) | 6<br>(0.6%) | 0<br>(0%) | 8<br>(0.8%) | 7<br>(0.7%) | 5<br>(0.5%) | 1<br>(0.1%) | 13<br>(1.3%) |
| Incomplete data | 23<br>(2.3%) | 101<br>(10%) | 51<br>(5.1%) | 175<br>(17.4%) | 14<br>(1.4%) | 106<br>(10.5%) | 54<br>(5.4%) | 174<br>(17.3%) |
| False data<br>("Hallucination") | 9<br>(0.9%) | 0<br>(0%) | 1<br>(0.1%) | 10<br>(1%) | 10<br>(1%) | 0<br>(0%) | 0<br>(0%) | 10<br>(1%) |
| Incorrect<br>calculations/inferences | 37<br>(3.7%) | 64<br>(6.4%) | 7<br>(0.7%) | 108<br>(10.7%) | 36<br>(3.6%) | 85<br>(8.4%) | 11<br>(1.1%) | 132<br>(13.1%) |
| Other | 0<br>(0%) | 5<br>(0.5%) | 1<br>(0.1%) | 6<br>(0.6%) | 2<br>(0.2%) | 8<br>(0.8%) | 0<br>(0%) | 10<br>(1%) |
| **Total** | **81**<br>**(8%)** | **343**<br>**(34.1%)** | **69**<br>**(6.9%)** | **493**<br>**(49%)** | **92**<br>**(9.1%)** | **333**<br>**(33.1%)** | **89**<br>**(8.8%)** | **514**<br>**(51%)** |

\* Percentages are rounded to the nearest one decimal point and thus total percentage might not be reflective of this.

### Costs

The overall costs of the AI-assisted and human-only arms were $3469 and $3651, respectively (Table 7 provides the cost breakdown).

**Table 7:** Breakdown of costs for AI-assisted and human-only data extraction arms.

| | AI-assisted arm (\$) | Human-only arm (\$) | Difference (\$) |
| --- | --- | --- | --- |
| Study file preparation<br>(Preparing data extraction forms and PDFs) | 75.23 | 75.23 | 0.00 |
| Study file preparation<br>(Uploading to Elicit®) | 56.17 | 0.00 | - 56.17 |
| Subscription to Elicit® <sup>+</sup> | 195.17 | 0.00 | - 195.17 |
| Elicit® preparation<br>(Prompt engineering and running Elicit®) | 756.29 | 0.00 | - 756.29 |
| Participants (Training and piloting) <sup>‡</sup> | 348.05 | 348.05 | 0.00 |
| Participants (Data extraction) | 2038.18 | 3227.78 | 1,189.61 |
| Total cost | 3469.09 | 3651.07 | 181.98 |

\$: Australian dollar; <sup>+</sup> Costs was adjusted to reflect actual days used; <sup>‡</sup> Participants did not train or pilot the use of Elicit®

Cost for each intervention arm included research assistant labour remuneration ^(45)^ (including data extraction, file preparation, setting up the AI-assisted arm, training) and annual Elicit^®^ Pro subscription fee ($763.70 per year) ^(37)^. The cost of Elicit^®^ Pro subscription was adjusted to reflect the actual days of use from the commencement of prompt engineering to the completion of data extraction.

## Discussion

To our knowledge, this is the first noninferiority SWAR examining the accuracy and efficiency of AI-assisted data extraction compared to human-only data extraction. The AI-assisted approach was noninferior and not significantly different in accuracy compared to the human-only approach, while making similar types and severity of errors – including hallucination rates. The AI-assisted approach was also noninferior. significantly faster and less costly. There were no differences in accuracy by data group and data type, with the exception of *intervention and control group extraction*, where AI-assisted data-extraction was significantly more accurate.

These findings are similar to other studies that utilised Elicit^®^ for data extraction ^(29, 33-35)^ and, more broadly, AI for data extraction ^(27, 30, 34, 53-60)^. While not directly comparable as conducted in different fields and employing different accuracy measures, previous evaluation of Elicit^®^ found an overall high precision, recall and F1-score of 92% for each measure ^(34)^.

We found that AI-assisted data extraction was significantly more accurate for ‘*intervention and control group’* data group than human-only data extraction. This finding aligns with Schmidt 2024, who reported high accuracy for ‘*intervention/control’* and *‘participants*’ data types ^(61)^. However, it contrasts with Gartlehner 2025, who found higher accuracy for ‘*participants’* and *‘study* characteristics’ ^(27)^, though neither study reported statistically significant differences. These discrepancies may be attributable to differences in the AI tools used (GPT-4 vs Claude 2 vs Elicit^®^). Future research is needed to explore these differences. For ‘*data and analysis’*, both arms in our trial scored approximately 75 points on a standard scale of 0-100, similar with Schmidt 2024, who reported low accuracy as well (<60%) ^(61)^. However another study reported 90.6% accuracy ^(27)^. This variation may be due to our use of single data extraction without consolidation or due to less experienced extractors, which likely contributed to the lower accuracy ^(4)^. These finding highlights that even if AI-assisted data extraction is noninferior to human-only extraction, more effort is needed to increase accuracy. Future research should explore if a consolidation process with AI could improve accuracy for *data and analysis*.

In terms of increasing efficiency, our findings indicate that the AI-assisted arm completed data extraction approximately 25 minutes faster per study compared to the human-only arm. This represents a substantial efficiency gain, particularly when scaled. For example, a typical systematic reviews includes approximately six studies ^(62, 63)^ to as many 195 studies ^(40)^, and if a similar AI-assisted data extraction process was applied, could translate to time savings of approximately 150 to 4875 mins. In the context of our trial, the AI-assisted arm collectively saved 1242 minutes in data extraction for 50 RCTs. Such time saving is important contributes to mitigating human fatigue (and possibly increasing error) and allows human experts to focus on other tasks that may be less suitable for AI-assistance such as evidence synthesis and interpretation ^(64)^.

Our trial confirms previous research ^(4, 11, 27)^, demonstrating that human-only data extraction is error-prone (10.1%), though mostly consisted of minor errors 6.5%, with the AI-assisted arm performing similarly (9.7% errors, 6.7% minor). Although concerns exist about AI data hallucinations, both arms showed an equally low rate of false data (0.2%), and the overall error patterns aligned with another LLM-based extraction study ^(27)^. It is important to note these findings are only applicable to AI-assisted data extraction tools which utilises human-in-the-loop, which ensures checks are in place to ensure the completeness of extracted information. While the findings of this trial are encouraging, a key concern regarding the use of LLMs in research should be noted. Specifically, LLMs, including those employed by Elicit^®^, lack transparency and replicability in their reasoning pathways and outputs, due to their inherently unobservable internal processes. Additionally, the rapid pace of AI development calls for careful consideration when integrating these technologies into systematic review workflows. A suggested way of continuing to integrate AI into data extraction processes is implementing a human-in-the-loop approach, as demonstrated in this trial, to ensure human oversight whilst improving efficiency ^(65)^. This approach can occur in several ways, such as employing two AI-assisted data extractors to maximize efficiency, or as demonstrated in this study, pairing one human with one AI-assisted extractor to balance efficiency with oversight, employing a second LLM to verify outputs or applying AI tools to assess study complexity and delegate difficult extractions to humans. Further exploration of such hybrid models is warranted, as another study proposed utilizing one human and one AI-only data extractor, followed by a second human to consolidate the two extractions ^(34)^. These varied methodologies highlight the substantial scope for continued research in this evolving field.

### Strengths and limitations

This trial has several key strengths. It is the first robust evaluation of Elicit^®^’s data extraction capabilities, assessing accuracy, time-to-complete, error types and severity, and cost within a simulated systematic review workflow. Another key strength is the use of a blinded assessor who verified each variable against the original papers using a predefined rubric, rather than relying on human-only extraction as the reference standard. This approach reduces benchmark bias, a bias when an imperfect reference standard is employed. Additionally, the large dataset of 5100 variable pairs ensures sufficient statistical power. Our prompt development process followed a systematic, transparent process, with all prompts publicly available for replication (Appendix B). Finally, the development of a three-point accuracy scale that captured partial correctness beyond binary measures offers a useful framework for future research.

Despite its strengths, our trial has limitations. First, reliance on an openly published review and publicly available trials as a data source, introduces a potential for data contamination, as Elicit^®^ may have been trained on this same data. Future research should integrate Elicit^®^ into a living systematic review to mitigate this risk and evaluate cost-effectiveness over multiple updates. Second, our findings are limited to the version of Elicit^®^ tested, with rapid advancements in LLM and newer features already available (e.g. all columns are now high-accuracy), future iterations may yield different and potentially improved results ^(34)^. Third, outcome data from both arms were unstructured and would require additional processing before analysis, which is an atypical workflow for a well-designed systematic review. However, this offers an opportunity for human oversight to ensure appropriate statistical considerations, such as standardising effects, units of measures and confirming adjustments for clustering, are addressed. Finally, this trial involved research assistants with relatively limited experience in systematic review data extraction, which may have influenced overall accuracy across both arms. Future studies could be undertaken with more experienced extractors (e.g. five years post-doctoral experience).

### Future directions

The promising findings from this study underscore the potential of AI for data extraction in systematic review workflows; however, further research is clearly warranted to explore different application scenarios. Future studies could investigate the efficacy of using two AI-assisted arms in parallel or assess a workflow where an AI tool checks the output of an AI-assisted arm before human oversight is applied. Furthermore, trials exploring other hybrid model approaches (e.g. using AI to assign difficult articles to humans) to replace human extractors would provide valuable insights. The findings from such investigations should then be rigorously incorporated into ongoing or living systematic reviews and directly compared against scenarios where AI was not utilised. This comparison would offer robust evidence of real-world applicability, foster greater trust of the use of AI, and potentially lead to clinically significant changes in practice.

## Conclusion

The use of Elicit^®^ is noninferior to human-only data extraction in both accuracy and time-to-complete. Both AI-assisted and human-only data extraction made similar types and severity of errors, and AI-assisted data extraction cost less than human-only data extraction. Future research should explore different application scenarios.

## Data Availability

All data produced in the present study are available upon reasonable request to the authors

## Acknowledgement

We would like to thank Ashley Blowes and Kate Mellors for participating in this trial.

## Author contributions

Study conception: DL, RH, KO, SY, JP, LW; Trial procedures: DL, KO; Statistical analysis: RH, CL; Interpretation of data: DL, RH, CL, AA, BH; Writing of manuscript: DL, RH, KO, SY, AA, JT, BH; Study supervision: RH, KO, JP, SY. All authors read and approved the final manuscript.

## Data management

Participation in the trial involved data extraction of research articles using Word Document, with no hard copy data recorded. All data were downloaded from OneDrive and stored in a confidential, de-identified file on a password-protected web server within the UON network, secured by a firewall. Access to identifiable data is restricted to the research team.

Data will be securely retained for a minimum of five years following research completion in accordance with the University’s Research Data and Materials Management Guideline (https://policies.newcastle.edu.au/document/view-current.php?id=72) or any successor guideline, and applicable UON policy provisions (as amended from time to time).

## Ethics and research approvals

Approval to conduct this trial to the methods described above was obtained from the University of Newcastle’s Human Research Ethics Committee, H-2024-0320. Elicit^®^ encrypts and does not allow uploaded PDFs to be accessed by any other user, which respects the intellectual property rights of copyrighted studies^(66)^.

